# Trends and Variation in User Fees across Provinces in China: a Population-based Longitudinal Data Analysis from 2011-2015

**DOI:** 10.1101/2020.06.15.20131813

**Authors:** Vicky Mengqi Qin, Yuting Zhang, Kee Seng Chia, Barbara McPake, Yang Zhao, Emily Hulse, Helena Legido-Quigley, John Tayu Lee

## Abstract

**Objectives:** Understanding the variation in user fees is essential for the design of targeted health financing strategies and monitoring progress towards universal health coverage. This study examines user fees in terms of: (1) temporal trends in cost sharing and out-of-pocket payment (OOPs); (2) factors associated with cost sharing and OOPs; and (3) the relationships between province-level economic development and cost sharing and OOPs in China.

**Setting:** 28 provinces of China

**Participants:** A total of 10316 elderly aged ≥45 years were included in the analysis.

**Results:** Overall, there were no substantial changes in cost sharing, but the OOPs continued to rise among the middle-aged and older Chinese. Cost sharing was substantially higher for outpatient than inpatient care (84.0% vs 69.2% in 2011; 80.8% vs 62.2% in 2015), and the majority paid more than 80% of the total cost for prescription drugs when visiting outpatient or inpatient care. Provinces with higher GDP per capita tend to have lower cost sharing and a higher OOPs than their counterparts, but the relationship for OOPs became insignificant after adjusting for individual factors. Respondents with health insurance and older age were associated with lower cost sharing. The respondents with higher socioeconomic status and a higher number of chronic conditions incurred higher OOPs for outpatient and inpatient utilisation.

**Conclusion:** Cost sharing and OOPs remain very high despite near-universal insurance coverage. Health financing reforms should prioritise improving health services coverage and reducing cost sharing to improve financial protection and reduce health inequalities. Although such improvement will likely have the greatest benefits for financial protection for populations from less developed regions, developed provinces with a higher OOPs will benefit as well.

**STRENGTHS AND LIMITATIONS OF THIS STUDY:**

- This is the first longitudinal study to measure the trend of and variation in patient cost sharing and OOPs in China.
- User fees was self-reported by the respondents, which may be subject to recall bias.
- User fees in this study only reflected the general cost burden from formal healthcare services, therefore, user fees from informal care services were not captured.

**KEY FINDING:**

- There were no substantial changes in patient cost sharing for outpatient and inpatient services, but the amount of out-of-pocket payment (OOPs) continued to rise during 2011-2015, especially for outpatient services from 371 Yuan in 2011 to 1031 Yuan in 2015.
- Despite universal health insurance coverage, patient cost sharing was still high among the middle-aged and older Chinese: 84.0% for outpatient care and 69.2% for inpatient care in 2011; and 80.8% vs 62.2% in 2015. The majority of patients paid more than 80% of the total cost for prescription drugs when visiting outpatient or inpatient care.
- Several patient-level characteristics affected cost-sharing and OOPs, including insurance status, age, education, household economic status and number of chronic conditions. Cost sharing was lower for those with insurance compared to those without insurance.
- Provinces with higher GDP per capita had lower cost sharing than provinces with lower GDP per capita, but no significant difference was found in the amount of OOPs after controlling for individual-level factors.

## INTRODUCTION

Protection against catastrophic levels of health spending as a result of illness has been a key goal of health systems in many countries.[1, 2] User fees, a direct payment at the point of seeking care paid by patients, remain the primary source of health care financing in many low-and middle-income countries.[3-6] Access to health care is inversely related to income and socioeconomic status, where wealthier groups have better access to high-quality health care than the poorer groups.[7, 8]

In China, almost the entire population (more than 95% in 2013) is covered by one of the three social health insurance schemes: the New Rural Cooperative Medical Scheme (NCMS), the Urban Resident Basic Medical Insurance (URBMI), and the Urban Employee Basic Medical Insurance (UEBMI). At the end of 2015, the Chinese government announced the decision to integrate URBMI and NCMS as the Urban-Rural Resident Medical Insurance Scheme. This integration has enabled a further extension of funding pools and narrowing disparities in access to health care services and medications that existed between different insurance schemes.[9] The three social health insurance schemes are designed to target different populations. The NCMS targets the registered rural population; the URBMI and UEBMI target the urban non-employed residents and employees, respectively. UEBMI generally provides more comprehensive service coverage (including both outpatient and inpatient services) and lower cost sharing compared with the other two schemes. [10] However, user fees for the same health insurance scheme can vary significantly across provinces due to fiscal capacity and priority setting of local governments.[11]

Despite the nearly universal health insurance coverage, as much as 13% of the population still face catastrophic payment in the recent years, indicating inadequate financial protection and a high level of user fees in China.[12] In addition, spending on medications has become a major component of total health expenditure (41.9% in 2010).[13] Thus, improving financial protection is crucial for health system strengthening in China, and the Chinese government has set an ambitious target to substantially reduce patient cost sharing (i.e. the percentage of out-of-pocket payment in total health expenditure) from 60% in 2001 to 25% by 2030.[14]

The literature on the provincial level variation in user fees is relatively sparse in China.[11, 15-17] A recent cost-sectional analysis of the key parameters of different health insurance programs found that cost sharing varies significantly by insurance schemes in China.[15] However, there is no longitudinal study to comprehensively document the individual and contextual factors associated with user fees and its changes over time, which could provide additional policy implication for future social health insurance reform. Using the longitudinal data from 2011-2015 of the China Health And Retirement Longitudinal Study (CHARLS), the present study examined: (1) trends in cost sharing and OOPs among middle-aged and older adults in China; (2) socioeconomic factors associated with user fees, and (3) the relationships between province-level economic development and user fees in China.

## METHODS

### Data

We used the longitudinal data from the China Health And Retirement Longitudinal Study (CHARLS) conducted in 2011, 2013, and 2015. CHARLS adopted the multi-stage stratified probability proportional to size sampling method at baseline. CHARLS had collected a nationally representative sample aged 45 years and above from 150 counties in 28 provinces. At baseline in 2011, 17,708 respondents (80.5% response rate) were interviewed, and 13,565 (76.6% of baseline sample) were followed up concurrently for three waves.[18] We identified 10,316 respondents, after removing ineligible respondents aged 45 years and below or with missing values in covariates.

### Measurements and variables

We measured user fees regarding: patient cost-sharing, defined as the ratio of out-of-pocket payment (OOPs) in total healthcare spending, and actual amount of OOPs (in Chinese Yuan).[19] We calculated them for outpatient and inpatient services separately. In addition, we also calculated user fees for prescription drugs which has been a major component of health spending in China. [13] We examined the association between socioeconomic determinants, geographic region, and user fees.

Respondents who sought outpatient care last month or inpatient care last year were asked: “*What was the total cost of this visit* (*or hospitalisation*), *including both treatment and medication cost* (*or fees paid to the hospital*)*?*”, and “*How much did you pay out of pocket, after reimbursement from insurance* (*for the total costs of hospitalisation*)*?*” Similarly, respondents were asked: “*What was the total medication cost for this visit, including prescription you received?*”, and “*How much will you eventually pay out of pocket for the medications from this visit, including prescriptions you received?*” for outpatient and inpatient settings, respectively. If there was no cost or respondents did not pay for the visits or medications, then patient cost sharing was denoted as 0 (i.e. partially compensated by insurance). Likewise, if the respondents further reported that the outpatient or inpatient visits were not covered by any insurance, then cost sharing was denoted as 1 (i.e. entirely paid by patients).

Socioeconomic indicators that may affect user fees were included in the analysis as independent variables, including: health insurance type (UEBMI, URBMI, NCMS, other insurance, without insurance), location (rural, urban), gender, age (45-54, 55-64, 65-74, ≥75), marital status (single/divorced/widowed, married/cohabitated), number of self-reported doctor diagnosed NCDs at individual level (none, 1 type of NCD, 2 types of NCDs, ≥ 3 types of NCDs), working status (working, retired, non-working), household economic status (the most deprived, deprived, middle, affluent and the most affluent), education (elementary school and below, secondary school, college and above) and time (year).[15, 20, 21] “Other insurance” included private insurance, government medical insurance (Gong Fei) and other supplementary insurance. We included 13 types of NCDs that are available in CHARLS for the calculation: hypertension, dyslipidaemia, diabetes, cancer, chronic lung disease, liver disease, heart disease, stroke, chronic kidney disease, digestive disease, mental disorders, arthritis and asthma. Household economic status was defined based on quintiles of yearly per capita household consumption. Per capita household consumption was based on relative approach by comparing yearly per capita household consumption to the median value at the city level to reduce bias from imbalance economic development across regions.[22] We also explored the relationship between user fees and economic development. We identified and ranked economic development at the provincial level based on their GDP per capita: low, <4300 US$; middle, 4300 −12000 US$; high, ≥12000 US$ (1 USD=6.2 CNY in 2014). The cut-off points for grouping provinces were referred to country classification from the World Bank.[23]

### Statistical analysis

We measured socioeconomic and provincial inequality in user fees using a series of regression-based methods.[24] We adopted a four-level random intercept linear regression model to explore the association between socioeconomic determinants and user fees, and to control for individual heterogeneity and measure external effects. OOPs was log-transformed in regression to allow for a more intuitive interpretation of regression coefficients as percentage changes in OOPs. The multilevel model accounted for hierarchical nature of the CHARLS data, with individuals at the first level, and community, city and province at the second, third and fourth level, respectively.

Therefore, cost sharing (denoted as Y_ijkl_) of individual i, living in community j of city k in province l in time t, given his/her sociodemographic characteristics can be described as follows:

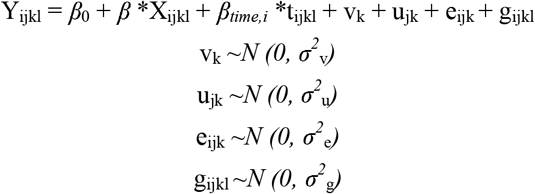

Where Y_ijkl_ is the predicted cost sharing, *β*_0_ is the mean cost sharing across participants, X_ijkl_ represents the vector of all independent variables that was adjusted for in the analysis with *β as the fixed effect, β*_*time,i*_ counted for the time effect, v_k_, u_jk_, e_ijk_ and g_ijkl_ represent the random effect of province, city, community, and individual respectively, assuming an independent and normal distribution with zero mean and constant variances (*σ*^*2*^_v_, *σ*^*2*^_u_, *σ*^*2*^_e_, *σ*^*2*^_g_).

We also measured the variance of cost sharing attributable to each level of the multilevel model by calculating variance partition coefficients (VPC). We excluded outliers with an extremely high value of health expenditures (i.e. >30,000 Yuan for outpatient, >300,000 Yuan for inpatient) before the calculation. To allow comparison over time, OOPs reported in 2011 and 2013 were converted to 2015 price using the gross domestic product (GDP) deflator according to the World Bank.[25] Adjusted coefficient (*β*) and 95% confidence intervals (CI) were presented for multilevel models, with p□<□0.05 taken as statistically significant. All statistical analyses were conducted using STATA 16.0.

## RESULTS

We analysed panel data from 10,316 respondents observed in 2011, 2013, and 2015. Table 1 summaries the sociodemographic characteristics of the respondents. At baseline, the majority of the respondents were female (51.1%), aged 55-64 years (38.9%), residing in rural areas (58.7%), currently working (71.4%), and attained elementary education or below (64.6%) in 2015. More than 65% of the respondents had at least one type of diagnosed NCD. More than 94% of participants were enrolled in at least one of the insurance schemes, with the majority insured by NCMS (73.7%).

**Table 1.**
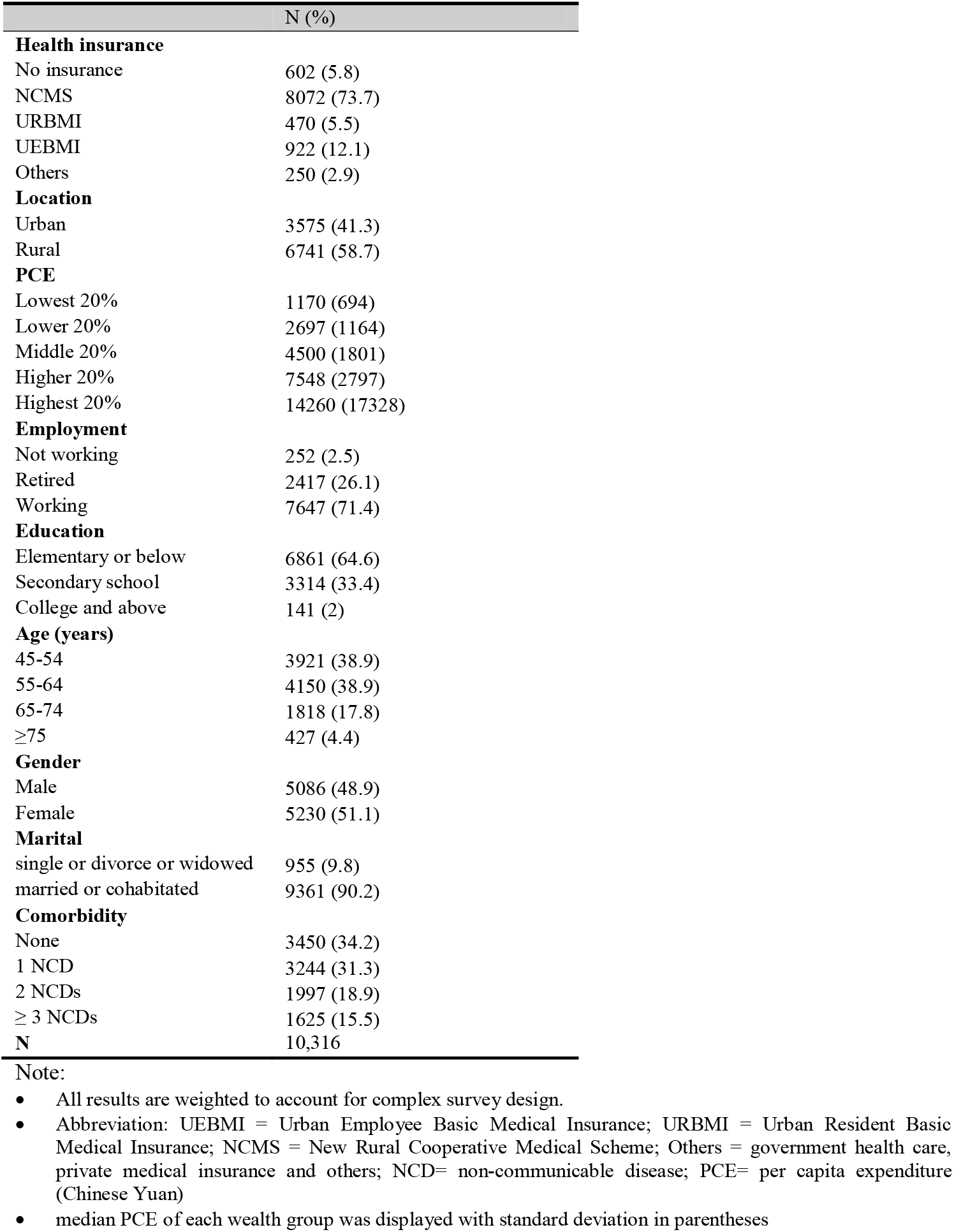
Sociodemographic characteristics of sample at baseline

### The trend in user fees 2011-2015

Overall, there was no substantial change in patient cost sharing between 2011-2015. However, the amount of OOPs continued to rise within the four year period (from an average of 371 Yuan to 1031 Yuan for outpatient, and from 4319 Yuan to 5952 Yuan for inpatient services).

Patient cost sharing was higher for outpatient than inpatient care (e.g. 84.0% vs 69.2% in 2011 and 80.8% vs 62.2% in 2015). The majority of the patients had to pay more than 80% of the total cost of prescription drugs when visiting outpatient or inpatient care.

Across the three major types of social health insurance schemes, participants enrolled in UEBMI had a lower cost sharing compared with participants enrolled in URBMI and NCMS. Participants enrolled in urban insurance (UEBMI or URBMI) had a higher OOPs than those insured by NCMS in rural areas. (Table 2)

**Table 2.**
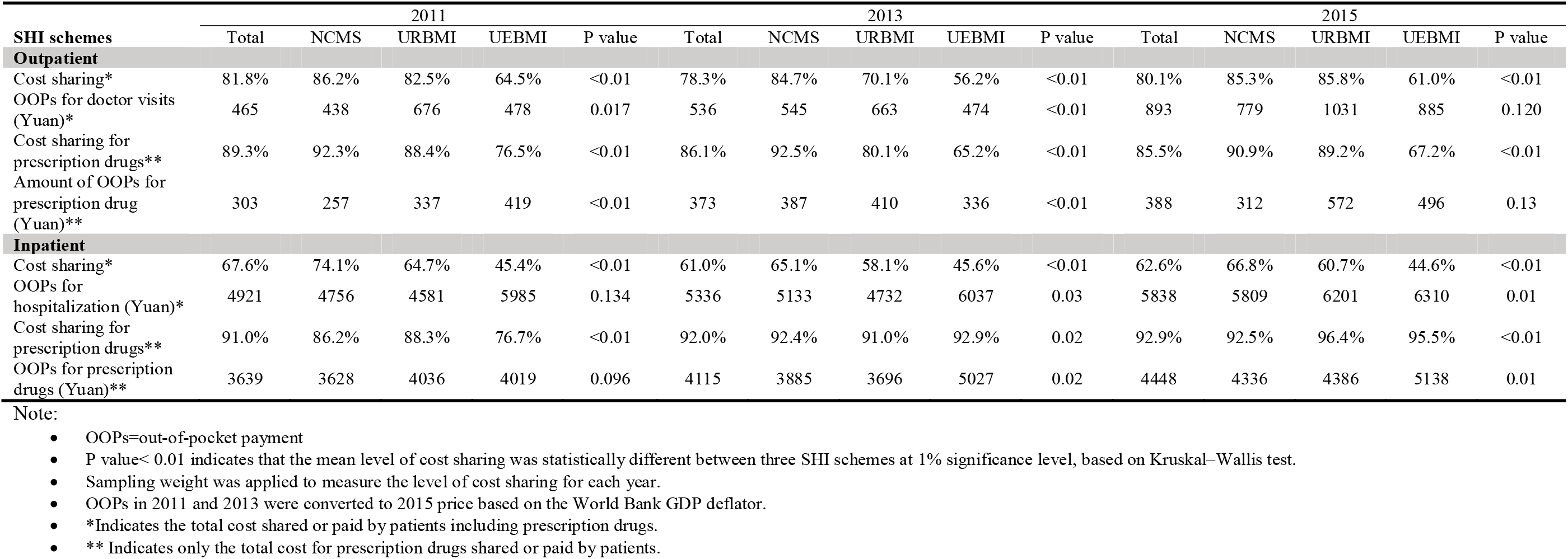
Patient Out-of-pocket Expenditure and Cost-sharing by Health Insurance schemes during 2011-2015

### Geographical variation

Provinces with higher GDP per capita tend to have lower cost sharing but higher OOPs for outpatient and inpatient care, compared with provinces with lower GDP per capita. Among provinces with high GDP per capita, the average cost sharing was 73.1% (1,341 Yuan) for outpatient visits, and 60.8% (7,641 Yuan) for inpatient visits in 2015. In comparison, the average cost sharing was 80.7% (579 Yuan) for outpatient visits, and 67.0% (4,505 Yuan) for inpatient visits among provinces with low GDP per capita.

### Percentage of cost sharing

#### Outpatient

Outpatient cost sharing was significantly lower among the insured respondents (regression coefficient=−0.09, −0.09, −0.21, −0.28, for NCMS, URBMI, UEBMI, and other health insurance respectively, p<0.05) than the uninsured counterparts (Table 3). People who were in the older age group (regression coefficient=−0.02, −0.05, −0.08, for those aged 55-64, 65-74, and 75 and above, p<0.05), retired (regression coefficient=−0.09, p<0.05), and had tertiary education (regression coefficient=−0.16, p<0.05) had lower cost sharing compared with those aged 45-54 years old, unemployed, and primary education or below, respectively. Outpatient cost sharing was lower among respondents from regions with high GDP per capita compared to respondents from regions with low GDP per capita (regression coefficient=−0.09, p<0.05). Outpatient cost sharing was not associated with gender, marital status, household economic status, and number of NCDs. There was no significant change of outpatient cost sharing during 2011 and 2015 (p>0.05).

**Table 3.** Determinants of patient cost sharing for outpatient and inpatient services from multilevel regression analysis

|  | Outpatient |  |  |  | Inpatient |  |  |  |
| --- | --- | --- | --- | --- | --- | --- | --- | --- |
|  | Overall |  | Medicines only |  | Overall |  | Medicines only |  |
|  | Regression coefficient | P value | Regression coefficient | P value | Regression coefficient | P value | Regression coefficient | P value |
| **Insurance type (ref: no insurance)** |  |  |  |  |  |  |  |  |
| NCMS | -0.089 (-0.127, -0.051) | <0.001 | -0.054(-0.086, -0.023) | 0.001 | -0.281(-0.334, -0.229) | <0.001 | -0.111(-0.164, -0.058) | <0.001 |
| URBMI | -0.094 (-0.144, -0.043) | <0.001 | -0.063(-0.105, -0.02) | 0.004 | -0.316(-0.382, -0.25) | <0.001 | -0.134(-0.2, -0.067) | <0.001 |
| UEBMI | -0.205 (-0.252, -0.158) | <0.001 | -0.171(-0.211, -0.131) | <0.001 | -0.453(-0.513, -0.393) | <0.001 | -0.172(-0.233, -0.111) | <0.001 |
| Others | -0.282 (-0.352, -0.211) | <0.001 | -0.278(-0.34, -0.215) | <0.001 | -0.508(-0.598, -0.418) | <0.001 | -0.167(-0.259, -0.076) | <0.001 |
| **PCE (ref: lowest 20%)** |  |  |  |  |  |  |  |  |
| Lower 20% | -0.022 (-0.048, 0.004) | 0.094 | 0.003(-0.019, 0.025) | 0.804 | 0.004(-0.036, 0.044) | 0.840 | -0.025(-0.065, 0.015) | 0.221 |
| Middle 20% | -0.025 (-0.05, 0) | 0.053 | -0.015(-0.036, 0.007) | 0.185 | -0.036(-0.074, 0.003) | 0.071 | -0.021(-0.06, 0.018) | 0.297 |
| Higher 20% | -0.015 (-0.041, 0.01) | 0.236 | -0.002(-0.023, 0.02) | 0.885 | 0.002(-0.035, 0.039) | 0.929 | -0.026(-0.064, 0.011) | 0.166 |
| Highest 20% | -0.02 (-0.046, 0.006) | 0.128 | -0.007(-0.029, 0.015) | 0.540 | 0.016(-0.02, 0.053) | 0.382 | -0.055(-0.092, -0.018) | 0.004 |
| **Employment status (ref: not working)** |  |  |  |  |  |  |  |  |
| Retired | -0.091 (-0.177, -0.005) | 0.038 | -0.045(-0.118, 0.028) | 0.228 | 0.056(-0.037, 0.15) | 0.236 | -0.033(-0.128, 0.062) | 0.495 |
| Working | -0.064 (-0.15, 0.021) | 0.140 | -0.026(-0.099, 0.047) | 0.488 | 0.046(-0.048, 0.14) | 0.335 | -0.031(-0.127, 0.064) | 0.518 |
| **Education (ref: primary school or below)** |  |  |  |  |  |  |  |  |
| Secondary school | -0.009 (-0.03, 0.011) | 0.369 | -0.004(-0.021, 0.013) | 0.652 | -0.028(-0.055, -0.002) | 0.036 | 0.002(-0.024, 0.029) | 0.866 |
| College and above | -0.164 (-0.24, -0.089) | <0.001 | -0.155(-0.221, -0.089) | <0.001 | 0.005(-0.086, 0.096) | 0.913 | -0.104(-0.197, -0.012) | 0.027 |
| **Age (ref: 45-54 years)** |  |  |  |  |  |  |  |  |
| 55-64 | -0.025 (-0.045, -0.005) | 0.013 | -0.013(-0.03, 0.004) | 0.125 | -0.031(-0.061, -0.002) | 0.035 | -0.012(-0.042, 0.017) | 0.411 |
| 65-74 | -0.054 (-0.078, -0.03) | <0.001 | -0.037(-0.058, -0.017) | <0.001 | -0.053(-0.086, -0.02) | 0.002 | -0.028(-0.061, 0.006) | 0.105 |
| 75 and above | -0.078 (-0.117, -0.04) | <0.001 | -0.074(-0.107, -0.04) | <0.001 | -0.082(-0.126, -0.037) | <0.001 | -0.053(-0.099, -0.008) | 0.022 |
| **Gender (ref: male)** |  |  |  |  |  |  |  |  |
| Female | 0.008 (-0.008, 0.025) | 0.324 | 0.019(0.004, 0.033) | 0.010 | 0.031(0.009, 0.053) | 0.006 | 0.031(0.009, 0.053) | 0.007 |
| **Marital (ref: single or divorce or widowed)** |  |  |  |  |  |  |  |  |
| Married or cohabitated | 0.004 (-0.019, 0.027) | 0.716 | -0.018(-0.037, 0.002) | 0.076 | 0.038(0.008, 0.069) | 0.014 | 0.004(-0.027, 0.035) | 0.784 |
| **Comorbidity (ref: none)** |  |  |  |  |  |  |  |  |
| 1 NCD | 0.004 (-0.023, 0.031) | 0.763 | 0.005(-0.019, 0.028) | 0.694 | 0.031(-0.01, 0.071) | 0.138 | -0.004(-0.045, 0.037) | 0.851 |
| 2 NCDs | 0 (-0.027, 0.027) | 0.995 | 0.011(-0.013, 0.034) | 0.364 | 0.015(-0.024, 0.054) | 0.452 | -0.008(-0.048, 0.032) | 0.682 |
| >2 NCDs | -0.005 (-0.032, 0.021) | 0.702 | 0(-0.023, 0.022) | 0.976 | 0.007(-0.03, 0.044) | 0.696 | -0.014(-0.051, 0.023) | 0.464 |
| **Location (ref: urban)** |  |  |  |  |  |  |  |  |
| Rural | 0.013 (-0.011, 0.038) | 0.290 | 0.003(-0.017, 0.022) | 0.774 | 0.045(0.016, 0.073) | 0.002 | -0.018(-0.046, 0.01) | 0.210 |
| **GDP per capita (ref: low)** |  |  |  |  |  |  |  |  |
| Middle | -0.015 (-0.043, 0.013) | 0.295 | -0.017(-0.04, 0.007) | 0.162 | 0.029(-0.007, 0.065) | 0.117 | -0.006(-0.04, 0.028) | 0.713 |
| High | -0.09 (-0.161, -0.019) | 0.013 | -0.057(-0.114, 0) | 0.050 | 0.064(-0.018, 0.145) | 0.128 | -0.002(-0.081, 0.076) | 0.953 |
| **Year (ref: 2011)** |  |  |  |  |  |  |  |  |
| 2013 | -0.019 (-0.041, 0.003) | 0.092 | -0.002(-0.021, 0.017) |  | -0.09(-0.122, -0.059) | <0.001 | 0.013(-0.018, 0.044) | 0.399 |
| 2015 | -0.011 (-0.035, 0.013) | 0.372 | -0.018(-0.039, 0.002) |  | -0.079(-0.112, -0.046) | <0.001 | 0.031(-0.001, 0.064) | 0.060 |
| **VPC, %** |  |  |  |  |  |  |  |  |
| Province | 1.5% |  | 0.6% |  | 0.0% |  | 0.0% |  |
| City | 1.6% |  | 1.6% |  | 2.7% |  | 0.5% |  |
| Community | 2.9% |  | 1.4% |  | 0.7% |  | 1.8% |  |
| Individual | 94.0% |  | 96.3% |  | 96.6% |  | 97.7% |  |

Note:
- 95% CI was displayed in parentheses after regression coefficient - Respondents categorized in other insurance group were enrolled in insurance program other than the three major social health insurance, such as private insurance, government funded insurance (Gong Fei) etc. - Abbreviation: UEBMI = Urban Employee Basic Medical Insurance; URBMI = Urban Resident Basic Medical Insurance; NCMS = New Rural Cooperative Medical Scheme; Others = government health care, private medical insurance and others; PCE= per capita expenditure (Chinese Yuan); NCD=non-communicable disease; VPC= Variance partition coefficient. - A likelihood ratio test was conducted to compare the fully adjusted model with null model with only random intercept and multivariable linear model. Fully adjusted multilevel model is preferred (P<0.01).

Likewise, people with insurance, tertiary education and older age also had lower cost sharing of prescription drugs in outpatient setting compared to their counterparts without insurance, low education level, and aged 45-54 years. Respondents from regions with high GDP per capita (regression coefficient=−0.05, p=0.05) had significantly lower cost sharing of prescription drugs than those from regions with low GDP per capita. Other sociodemographic covariates such as household economic status, employment status, and number of NCDs were not associated with outpatient cost sharing of prescription drugs (p>0.05).

#### Inpatient

Inpatient cost sharing was significantly lower among respondents with health insurance (regression coefficient=−0.28, −0.32, −0.45, −0.51, for NCMS, URBMI, UEBMI, and other health insurance respectively, p<0.05). Respondents who were female (regression coefficient=0.03, p<0.05), married (regression coefficient=0.04, p<0.05), and resided in rural area (regression coefficient=0.05, p<0.05) had higher level of cost sharing than their counterparts. Respondents who were aged between 55-64, 65-74, and 75 and older had lower level of cost sharing (regression coefficient=− 0.03, −0.05, −0.08, p<0.05), compared with those aged 45-54 years. The level of inpatient cost sharing was lower in year 2013 (regression coefficient= −0.10, p<0.05) and 2015 (regression coefficient=−0.09, p<0.05), compared with year 2011. Cost sharing for inpatient services was not significantly associated with education level, employment status, household economic status, number of NCDs, and regional economic development.

Similarly, inpatient cost sharing of prescription drugs was lower among people with insurance, aged 75 and above, had tertiary education compared to those without insurance, aged 45-54, and had primary education or below, respectively. Inpatient cost sharing of prescription drugs was not significantly different with regard to employment status, household economic status, location and provincial economic development (p>0.05).

### Amount of OOPs

#### Outpatient

Table 4 shows that the amount of OOPs for outpatient visits was lower among respondents insured by UEBMI (regression coefficient=−0.34, p<0.05) and “other insurance” including private and government funded insurance (regression coefficient=−1.17, p<0.05) compared to those without insurance. Older age groups (regression coefficient=−0.13, −0.21 and −0.48 for age group 55-64, 65-74 and 75 and above respectively, p<0.05) and tertiary education (regression coefficient=−0.80, p<0.05) was associated with less OOPs, compared to people aged 45-54 years and who had primary education or below respectively. Respondents from households with the most affluent economic status spent more for outpatient OOPs compared to those from the worst economic status (regression coefficient=0.39, p<0.05). Respondents who were married (coefficient=0.21, p<0.05) and had more NCDs (coefficient=0.27, 0.42, 0.46, for people had two and more than two types of NCDs, p<0.05) spent higher OOPs than their counterparts who were male, single, and without diagnosed NCDs respectively. OOPs was higher in year 2013 (regression coefficient= 0.21, p<0.05) and 2015 (regression coefficient=0.45, p<0.05), compared with year 2011. Outpatient OOPs were also not associated with employment status, education, location, and regional economic development.

**Table 4.** Determinants of the amount of OOPs for outpatient and inpatient services from multilevel analysis

|  | Outpatient |  |  |  | Inpatient |  |  |  |
| --- | --- | --- | --- | --- | --- | --- | --- | --- |
|  | Log of OOPs |  | Log of OOPs for Prescription drugs |  | Log of OOPs |  | Log of OOPs for Prescription drugs |  |
|  | Regression coefficient | P value | Regression coefficient | P value | Regression coefficient | P value | Regression coefficient | P value |
| **Insurance type (ref: no insurance)** |  |  |  |  |  |  |  |  |
| NCMS | -0.094(-0.339, 0.152) | 0.455 | -0.071(-0.295, 0.152) | 0.532 | -0.08(-0.427, 0.267) | 0.652 | -0.065(-0.443, 0.314) | 0.738 |
| URBMI | 0.012(-0.312, 0.337) | 0.941 | 0.111(-0.186, 0.408) | 0.465 | -0.021(-0.454, 0.412) | 0.925 | -0.065(-0.538, 0.408) | 0.788 |
| UEBMI | -0.338(-0.642, -0.035) | 0.029 | -0.266(-0.544, 0.011) | 0.060 | -0.002(-0.401, 0.398) | 0.993 | 0.035(-0.4, 0.469) | 0.876 |
| Others | -1.173(-1.626, -0.72) | <0.001 | -1.333(-1.761, -0.905) | <0.001 | -1.101(-1.686, -0.516) | <0.001 | -0.347(-0.991, 0.298) | 0.292 |
| **PCE (ref: lowest 20%)** |  |  |  |  |  |  |  |  |
| Lower 20% | -0.141(-0.306, 0.024) | 0.093 | -0.048(-0.199, 0.104) | 0.538 | 0.115(-0.14, 0.371) | 0.376 | -0.085(-0.366, 0.197) | 0.556 |
| Middle 20% | 0.092(-0.07, 0.254) | 0.264 | 0.081(-0.067, 0.23) | 0.283 | 0.182(-0.066, 0.43) | 0.151 | 0.097(-0.177, 0.372) | 0.486 |
| Higher 20% | 0.112(-0.051, 0.275) | 0.178 | 0.131(-0.018, 0.28) | 0.085 | 0.493(0.255, 0.732) | <0.001 | 0.222(-0.04, 0.484) | 0.097 |
| Highest 20% | 0.391(0.226, 0.556) | <0.001 | 0.361(0.21, 0.512) | <0.001 | 0.759(0.523, 0.995) | <0.001 | 0.326(0.066, 0.586) | 0.014 |
| **Employment status (ref: not working)** |  |  |  |  |  |  |  |  |
| Retired | -0.156(-0.712, 0.4) | 0.582 | -0.102(-0.611, 0.408) | 0.696 | 0.202(-0.402, 0.806) | 0.513 | -0.466(-1.127, 0.195) | 0.167 |
| Working | -0.444(-0.998, 0.109) | 0.116 | -0.407(-0.915, 0.1) | 0.116 | -0.158(-0.765, 0.448) | 0.608 | -0.739(-1.401, -0.077) | 0.029 |
| **Education (ref: primary school or below)** |  |  |  |  |  |  |  |  |
| Secondary school | -0.001(-0.13, 0.128) | 0.988 | 0.011(-0.107, 0.129) | 0.859 | -0.096(-0.271, 0.079) | 0.282 | 0.071(-0.115, 0.257) | 0.456 |
| College and above | -0.802(-1.275, -0.328) | 0.001 | -0.488(-0.941, -0.036) | 0.035 | -0.056(-0.654, 0.541) | 0.853 | -0.78(-1.42, -0.14) | 0.017 |
| **Age (ref: 45-54 years)** |  |  |  |  |  |  |  |  |
| 55-64 | -0.13(-0.257, -0.002) | 0.046 | -0.108(-0.224, 0.008) | 0.069 | -0.163(-0.354, 0.027) | 0.093 | -0.162(-0.368, 0.043) | 0.121 |
| 65-74 | -0.208(-0.361, -0.056) | 0.008 | -0.145(-0.285, -0.005) | 0.042 | -0.425(-0.64, -0.21) | <0.001 | -0.356(-0.588, -0.124) | 0.003 |
| 75 and above | -0.482(-0.727, -0.237) | <0.001 | -0.381(-0.608, -0.154) | 0.001 | -0.74(-1.033, -0.447) | <0.001 | -0.498(-0.814, -0.181) | 0.002 |
| **Gender (ref: male)** |  |  |  |  |  |  |  |  |
| Female | 0.036(-0.071, 0.143) | 0.508 | 0.021(-0.077, 0.119) | 0.680 | -0.129(-0.273, 0.016) | 0.081 | -0.106(-0.261, 0.048) | 0.177 |
| **Marital (ref: single or divorce or widowed)** |  |  |  |  |  |  |  |  |
| Married or cohabitated | 0.212(0.066, 0.357) | 0.004 | 0.075(-0.059, 0.209) | 0.273 | 0.482(0.283, 0.681) | <0.001 | 0.145(-0.069, 0.359) | 0.185 |
| **Comorbidity (ref: none)** |  |  |  |  |  |  |  |  |
| 1 NCD | 0.268(0.095, 0.44) | 0.002 | 0.273(0.115, 0.432) | 0.001 | 0.159(-0.103, 0.421) | 0.233 | 0.114(-0.172, 0.4) | 0.435 |
| 2 NCDs | 0.417(0.243, 0.59) | <0.001 | 0.425(0.265, 0.585) | <0.001 | 0.071(-0.184, 0.326) | 0.584 | -0.005(-0.283, 0.272) | 0.971 |
| >2 NCDs | 0.46(0.291, 0.628) | <0.001 | 0.431(0.276, 0.586) | <0.001 | 0.129(-0.111, 0.369) | 0.291 | 0.021(-0.24, 0.281) | 0.877 |
| **Location (ref: urban)** |  |  |  |  |  |  |  |  |
| Rural | 0.091(-0.061, 0.243) | 0.240 | 0.049(-0.084, 0.183) | 0.469 | 0.001(-0.184, 0.187) | 0.988 | -0.349(-0.554, -0.144) | 0.001 |
| **GDP per capita (ref: low)** |  |  |  |  |  |  |  |  |
| Middle | -0.131(-0.312, 0.05) | 0.157 | -0.104(-0.273, 0.064) | 0.225 | 0.236(-0.004, 0.476) | 0.054 | 0.146(-0.103, 0.395) | 0.251 |
| High | -0.208(-0.669, 0.254) | 0.378 | -0.064(-0.498, 0.371) | 0.773 | 0.649(0.094, 1.204) | 0.022 | 0.605(0.011, 1.2) | 0.046 |
| **Year (ref: 2011)** |  |  |  |  |  |  |  |  |
| 2013 | 0.212(0.07, 0.354) | 0.003 | 0.137(0.006, 0.269) | 0.041 | -0.096(-0.297, 0.104) | 0.347 | 0.049(-0.174, 0.272) | 0.664 |
| 2015 | 0.447(0.292, 0.602) | <0.001 | 0.155(0.011, 0.299) | 0.035 | -0.039(-0.252, 0.175) | 0.723 | 0.136(-0.1, 0.371) | 0.258 |
| **VPC, %** |  |  |  |  |  |  |  |  |
| Province | 2.0% |  | 3.0% |  | 1.1% |  | 0.3% |  |
| City | 2.3% |  | 2.4% |  | 0.6% |  | 0.0% |  |
| Community | 1.8% |  | 1.1% |  | 1.9% |  | 3.8% |  |
| Individual | 93.9% |  | 93.5% |  | 96.4% |  | 95.9% |  |

Note:
- 95% CI was displayed in parentheses after regression coefficient - Respondents categorized in other insurance group were enrolled in insurance program other than the three major social health insurance, such as private insurance, government funded insurance (Gong Fei) etc. - Abbreviation: UEBMI = Urban Employee Basic Medical Insurance; URBMI = Urban Resident Basic Medical Insurance; NCMS = New Rural Cooperative Medical Scheme; Others = government health care, private medical insurance and others; OOPs=out-of-pocket payment; PCE= per capita expenditure (Chinese Yuan); VPC= Variance partition coefficient. - A likelihood ratio test was conducted to compare the fully adjusted model with null model with only random intercept and multivariable linear model. Fully adjusted multilevel model is preferred (P<0.01). - OOPs was log transformed to normalize the distribution. Patients with zero OOPs were replaced by 1 for a mathematically meaningful log-transformation.

Outpatient OOPs of prescription drugs were also lower among respondents insured by UEBMI and “other insurance”, being in older age group groups than the uninsured group and those aged 45-54 years. Respondents from the most affluent households (coefficient=0.36, p<0.05) and who had more NCDs (coefficient=0.43, for people who had two and more than two types of NCDs, p<0.05) spent a higher amount of OOPs compared their counterparts. No significant difference was found for outpatient OOPs of prescription drugs with regards to gender, marital status, employment status, location, and provincial economic development.

#### Inpatient

OOPs for inpatient services did not significantly differ by the type of social health insurance compared to those without any insurance. However, people covered by “other insurance” such as private insurance (regression coefficient=−1.10, p<0.05) spent less OOPs than those without insurance. People in the older age groups spent less for inpatient OOPs compared to those aged 45-54 years (regression coefficient=− 0.43, −0.74, for 65-74 and 75 and above, respectively, p<0.05). Respondents who were married (regression coefficient=0.48, p<0.05) and had the most affluent household economic status (regression coefficient=0.76, p<0.05) spent more on inpatient OOPs compared to those single and with the most deprived household economic status. Respondents from provinces with middle and high GDP per capita spent more on OOPs (regression coefficient=0.24, 0.65, for middle and high GDP per capita respectively p<0.05) compared to those from provinces with low GDP per capita. Inpatient OOPs were not associated with gender, employment status, education, number of NCDs, and location.

Respondents who were in the older age group (regression coefficient=−0.35, −0.49, for aged 65-74 and 75 and above respectively, p<0.05), employed (regression coefficient=−0.74, p<0.05), and had tertiary education (regression coefficient=−0.80, p<0.05) spent less on OOPs for prescription drugs during hospitalisation, compared to those aged 45-54 years and unemployed, respectively. People from the most affluent household (regression coefficient=0.33, p<0.05) spent more on OOPs for prescription drugs than their counterparts from the most deprived household. OOPs for prescription drugs was significantly higher among respondents from regions with high GDP per capita compared to respondents from regions with low GDP per capita (regression coefficient=0.61, p<0.05). Inpatient OOPs for prescription drugs was not associated with gender, number of NCDs, and location.

#### Partitioning variations in user fees

In the fully adjusted model for cost sharing, 1.5% of the variation in outpatient costsharing comes from provinces, 1.6% from cities, 2.9% from communities within cities, and 94% lies within the community between individuals (Table 3). In inpatient settings, individuals accounted for 96.6% of the variation in cost sharing, followed by 0.7% and 2.7% at the community and city-level respectively.

Variation in outpatient OOPs was similar, with individuals accounted for 93.9% of the variation, followed by communities (1.8%), cities (2.3%) and provinces (2%). Individual-level accounted for 96.4% of variation in inpatient OOPs, with community, city and province-level accounted for 1.9%, 0.6% and 1.1%. (Table 4).

## DISCUSSION

### Principal findings

Using the longitudinal data from 10,316 respondents aged 45 years and above in China, we found no substantial change in the percentage of cost sharing over time, but the amount of OOPs continued to rise. Cost sharing and OOPs were lower among insured than the uninsured group, UEBMI than the other social health insurance schemes, and private insurance than the social health insurance schemes. Provinces with higher GDP per capita (such as Beijing, Shanghai and Tianjin) tend to have lower cost sharing but higher OOPs than those provinces with lower GDP per capita (such as Yunnan, Guizhou, Gansu). Covered by health insurance scheme was associated with lower cost sharing and less OOPs. Collectively, these results suggest that the uninsured population from the less developed provinces with worse health conditions are at greatest financial burden due to illness in China.

### Interpretation

This is the first study that adopted robust longitudinal study design to examine differences in user fees across provinces, types of health insurance, and socio-demographic groups in China. Consistent with previous studies, we found that the type of health insurance coverage is a significant determinant of cost sharing after controlling for socioeconomic factors.[15, 17, 26, 27] Compared with urban insurance schemes, people with NCMS or without insurance were at greater risk of financial burden. Fragmented and low SHI benefit coverage may be due to the low premiums contribution (especially in the rural insurance scheme) and restricted spending of social health insurance funds (consists of less than 36% of total health expenditure with a surplus rate of more than 25% in urban insurance scheme by 2016).[27-29].

Our findings showing patient cost sharing was higher for the outpatient than inpatient visits, which is consistent with previous studies as well as the report from CHARLS dataset.[30, 31] This indicates that social health insurance has put more emphasis on inpatient than outpatient services. We found that people from households with better economic status incurred a higher amount of OOPs from seeking treatment for their illness, which is similar to the conclusion from previous studies based on cross-sectional data.[15, 17] This is likely because those economically better-off households have a higher intensity of health care use and received health care from higher-level health care provider (such as a tertiary hospital).[33]

The results from the unadjusted models reveal that provinces with higher GDP per capita tended to have lower cost sharing but higher amount of OOPs. The association become insignificant for cost sharing in inpatient setting and OOPs in outpatient setting after controlling for other covariates. These findings were in general consistent with an earlier study.[11] Recent health reform has been focusing on broadening insurance coverage to include services that were previously not covered.[36] Provinces with advanced economic development may have prioritised or had a larger investment in the health sector which enhances financial protection by lowering patient cost sharing.[37] On the other hand, higher OOPs in wealthier provinces could be due to the more expensive health services used that were not covered by insurance, or the dominance of secondary and tertiary health facilities.

### Strengths and limitations

To our knowledge, this is the first longitudinal study to measure the trend of and variation in patient cost sharing and OOPs in China. A multilevel modelling method using longitudinal data was adopted to obtain robust results. Our study had several limitations. Firstly, user fees were self-reported by the respondents, which may be subject to recall bias due to inaccurate or incomplete reporting, particularly with older participants. Secondly, user fees in this study measured the general cost burden for seeking health care. Therefore, user fees for specific types of disease was not available. Thirdly, data on expenditure was only available among the elderly who visited outpatient care last month or inpatient care last year. It is possible that those who did not seek care have better health conditions and face different user fees when seeking care. Generalisability of the results might also be exclusive of people aged 45 years and below who may have different patterns and determinants of user fees. Lastly, our findings of user fees only reflect the level of financial protection from formal healthcare services (such as hospital and clinics). Therefore, user fees from informal care (such as purchasing medicines over the counter) was not captured and beyond the scope of this study.[38, 39]

### Policy implications

Despite the nearly universal health insurance coverage in China, our findings provide more evidence for health insurance reform to address the high level of and the large variation in user fees. Moving towards universal health coverage in China needs to address the issue of disparity in accessing health care, of which reducing user charge is critical.[40] Although a few provinces have benefited from relatively low cost sharing, high cost sharing (higher than 30%, a threshold considered as low level of financial protection) and increasing OOPs in most provinces, especially people from rural regions, should not be neglected.[10]

We also found that the percentage of cost sharing for prescription drugs remained high nationwide. The high cost sharing for outpatient raises a concern for an aging population who need longer-term access to outpatient care and medication treatment with an increasing prevalence of NCDs. Recent evidence shows that the introduction of a reimbursement for the outpatient cost of NCDs in NCMS has yet to reduce the incidence of CHE effectively.[41] Hence, enhancing financial support of social health insurance should continuously target lowering cost-sharing for long-term/chronic prescription drugs for people with NCDs/chronic diseases.[32] Although the national essential drug policy reform reduced drug expenditure at inpatient settings, it has yet to significantly reduce drug expenditure at outpatient settings and for total health spending.[42, 43] Policies to broaden the benefits package of social health insurance, such as expanding the essential medicines list, should be prioritised.[44] While policies to lower mark-up for prescription drugs can reduce price, complementary financing mechanism such as government subsidies should be considered to counteract the income loss suffered by healthcare facilities and providers from drug sales.[13] Effectively lowering cost sharing is an essential strategy to make the needed health services accessible and affordable, that could reduce inequalities and yield more substantial and sustainable impacts on overall population health.[45, 46]

Reducing patient user fees alone is not sufficient to improve protection against financial risk. A broader and systemic health system reform is needed to maintain the sustainability of health financing mechanisms.[47, 48] This should include improvements in quality and efficiency of primary care as well as targeted cost sharing design to ensure less reliance on unnecessary secondary and tertiary care.[10] Future research is needed to evaluate the impact of these new initiatives implemented and to examine the effect of reimbursement intervention that reduced cost-sharing in China.[32]

## Conclusion

Patient cost sharing and OOPs remain high and provincial and socioeconomic variation exists between 2011-2015, despite near-universal insurance coverage. Improving health services coverage is needed to reduce user fees and narrow regional inequality. Although such improvement will likely have the greatest benefits for financial protection for populations from less developed regions, developed provinces with a higher OOPs will benefit as well.

**Figure 1.**
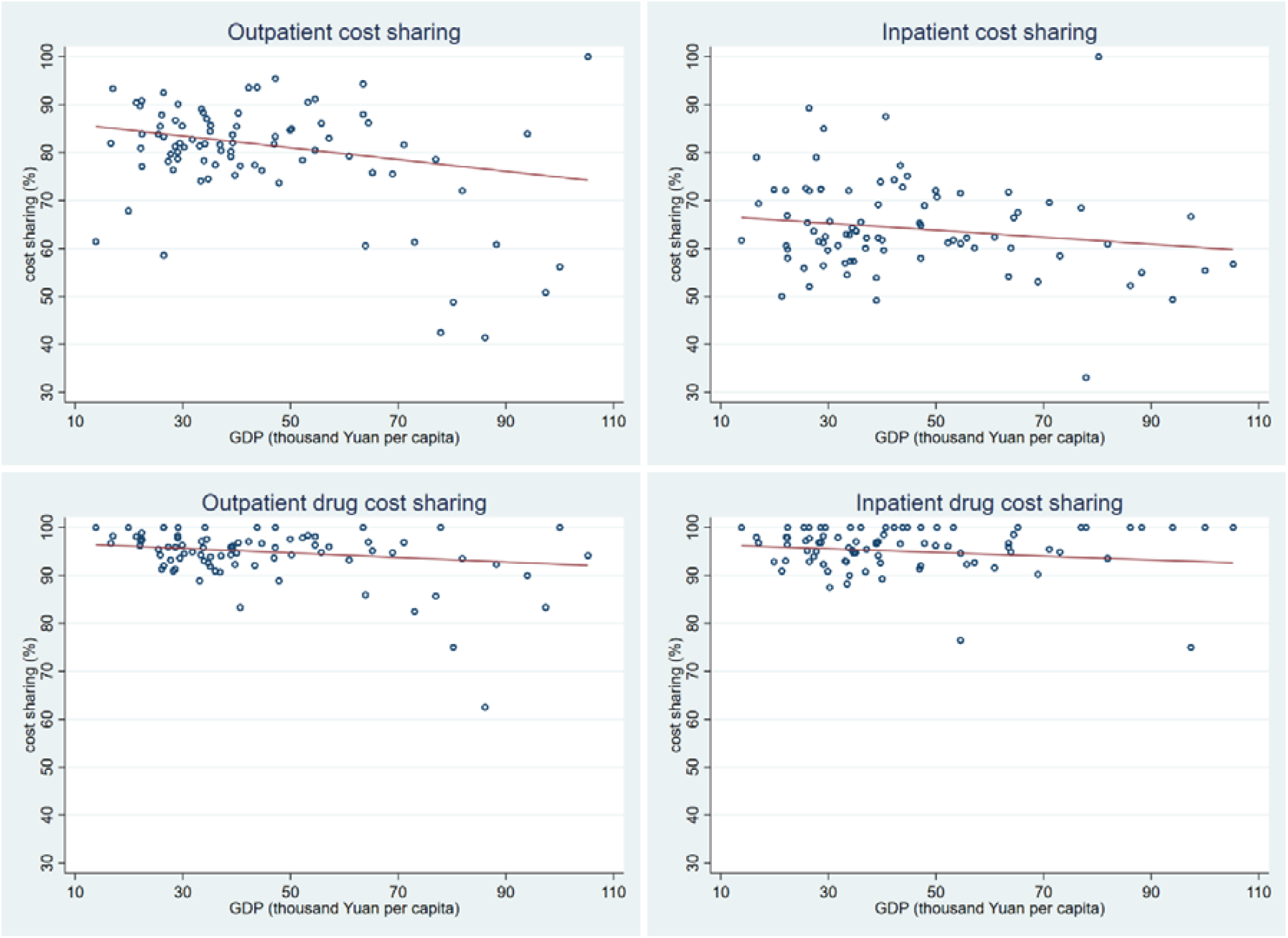
Bivariate relationship between cost sharing and economic development at provincial level between 2011-2015 (n=84) Note: Provincial GDP per capita data was extracted from China Statistical Yearbook 2016. Each province (n=28) has three independent data points.

**Figure 2.**
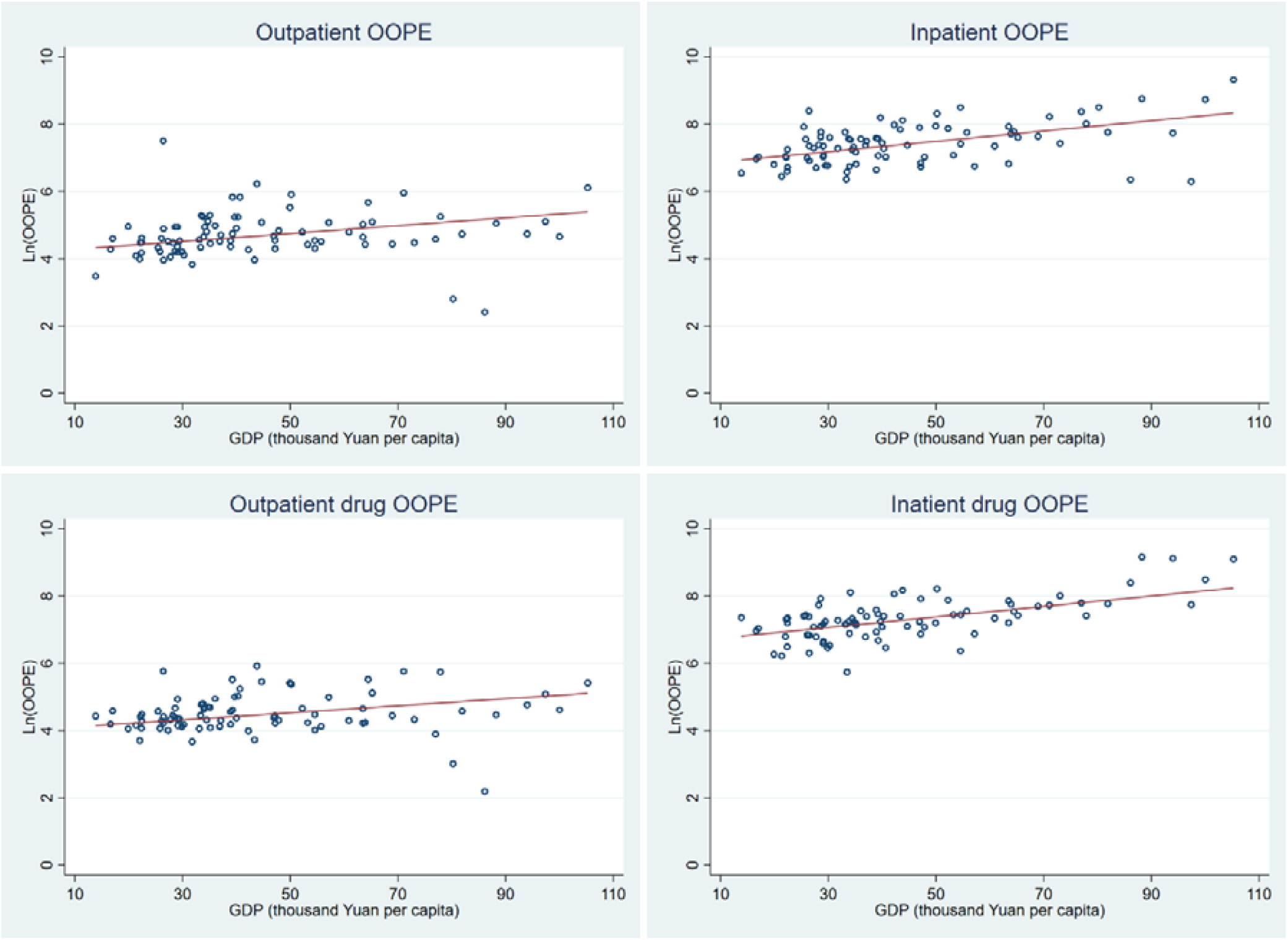
Bivariate relationship between OOPs and economic development at provincial level between 2011-2015 (n=84) Note: Provincial GDP per capita data was extracted from China Statistical Yearbook 2016. OOPs was log transformed to normalize the distribution. OOPs in 2011 and 2013 were converted to 2015 price-based GDP deflator. Each province (n=28) has three observations.

## Data Availability

Data is publicly available and can be found via http://charls.pku.edu.cn/index/en.html

## ABBREVIATION

GDP: gross domestic product
NCD: non-communicable diseases
NCMS: new cooperative medical scheme
OOPs: out-of-pocket payment
UEBMI: urban employee basic medical insurance
URBMI: urban resident basic medical insurance

## Funding

This research received no specific grant from any funding agency in the public, commercial or not-for-profit sectors.

## Contributors

VMQ and JTL conceived the study. VMQ conducted the data analysis and wrote the manuscript. YTZ, EH and JTL helped on editing the manuscript. YTZ, KSC, BM, YZ, HL and JTL critically commented on the revision and approved the final version.

## Patient consent form

CHARLS was approved by the Ethical Review Committee (IRB) at Peking University, Beijing, China. All participants were required to provide written informed consent.

## A data sharing statement

None.

## Conflict of interest

None declared.

## Notes

### Competing Interest Statement

The authors have declared no competing interest.

### Author Declarations

CHARLS was approved by the Ethical Review Committee (IRB) at Peking University, Beijing, China.

